# Prevalence and determinants of non-communicable diseases risk factors among reproductive-aged women of Bangladesh: Evidence from Bangladesh Demographic Health Survey 2017-2018

**DOI:** 10.1101/2022.08.03.22278387

**Authors:** Saifur Rahman Chowdhury, Md Nazrul Islam, Tasbeen Akhtar Sheekha, Shirmin Bintay Kader, Ahmed Hossain

## Abstract

**Introduction:** Knowing the risk factors like hypertension, overweight/obesity, and smoking status among women of reproductive age could allow the development of an effective strategy for reducing the burden of non-noncommunicable diseases (NCD). We sought to determine the prevalence and determinants of NCD risk factors among Bangladeshi women of reproductive age.

**Methods:** This study utilized the Bangladesh Demographic and Health Survey (BDHS) data from 2017-2018 and analyzed 5,624 women of reproductive age. This nationally representative cross-sectional survey utilized a stratified, two-stage sample of households. Mixed-effects Poisson regression models were fitted to find the adjusted prevalence ratio for smoking, overweight and obesity, and hypertension.

**Results:** The average age of 5,624 participants was 31 years (SD=9.07). The prevalence of smoking, overweight/obesity, and hypertension was 9.55%, 31.57%, and 20.27%, respectively. More than one-third of the participants (34.55%) had one NCD risk factor, and 12.51% of participants had two NCD risk factors. Women between 40–49 years had more NCD risk factors than 18–29 years aged women (APR: 2.44; 95% CI: 2.22-2.68). Women with no education (APR: 1.15; 95% CI: 1.00-1.33), married (APR: 2.32; 95% CI: 1.78–3.04), and widowed/divorced (APR: 2.14; 95% CI: 1.59–2.89) were more prevalent in NCD risk factors. Individuals in the Barishal (APR: 1.44; 95% CI: 1.28–1.63) division were living with higher risk factors for NCD. Women who belonged to the richest (APR: 1.82; 95% CI: 1.60–2.07) wealth quintile were more likely to have the risk factors of NCD.

**Conclusions:** This study revealed that older women, currently married and widowed/divorced women, women from the wealthiest socio-economic group, and women with a lower level of education were more likely to have NCD risk factors. Furthermore, there was a significant relationship between the geographical division and NCD risk factors. To reduce the future prevalence of NCD, it is necessary to implement effective prevention and control programs that target women with a higher risk of having the disease.

## Introduction

The major cause of global death and disability is non-communicable diseases (NCD) [1,2]. The most common NCD are cardiovascular diseases (CVD), cancer, chronic respiratory diseases, and diabetes [2]. Every year an estimated 85% of 15 million premature deaths occur due to NCD in low- and middle-income countries (LMICs) worldwide [2], and almost 80% of the deaths occur due to the above-mentioned NCD [2]. Between 2012 and 2030, the number of deaths due to NCD is predicted to reach 52 million from 38 million globally [3].

In Bangladesh, NCD are significant cause of mortality, responsible for 67% of total deaths [4]. The prevalence of NCD has increased over the last ten years, and it is predicted to increase more as Bangladesh is passing through a growing stage of epidemiological transition [1,5–7]. Bangladesh is a lower-middle-income country having over 170 million people [8]. It surfaced as a developing country with an experience of rapid growth in economy and urbanization in the previous decades [8]. As a result of this substantial development, there is a large number of people are leading a more sedentary lifestyle influenced by the change in dietary habits, increased supply and demand for unhealthy processed food, and physical inactivity followed by irregularities in mealtimes, smoking, and alcohol consumption [7–10]. Moreover, there are metabolic risk factors that contribute to the development of NCD. The most common metabolic risk factors are obesity, high blood sugar, high blood pressure, and increased cholesterol [6]. There is an increased chance of developing NCD when two or more of these factors combine, referred to as clustering of risk factors [11]. Consequently, a considerable proportion of the population suffered from overweight (29.4%) [12], hypertension (26.2%) [13], and diabetes (9.2%) [14] in Bangladesh.

Women are at an increased risk of developing NCD as they mostly experience a combination of multiple behavioral and metabolic risk factors [15,16]. Globally, NCD are responsible for 65% of premature death in women, with the majority of death occurring in LMICs [17]. Evidence showed that between 1992 and 2015, the prevalence of diabetes has a threefold to fourfold increase, with a prediction to reach around 24% in men and 33.5% in women by 2030, with higher odds of multimorbidity than men [18]. The reason behind this is that women having low socio-economic conditions than men causing gender inequality which affects health outcomes [19]. The fetal health and the reproductive health of women become severely affected by the NCD risk factors [20,21]. For example, hypertension in pregnancy increases menstrual problems, complications in pre- and post-natal periods, and maternal mortality [22]. Moreover, previous research showed that women were more likely to be overweight in urban areas because of rapid urbanization, whereas they were more likely to be underweight in rural areas because of inadequate provision of healthy food [23]. Lack of proper nutrition may lead to serious metabolic changes contributing to the development of heart disease, hypertension, and diabetes among them as well as among their future generation [9]. In addition, obesity is related to polycystic ovary syndrome (PCOS) [24], and hypertension is the common cause of higher maternal mortality and stillbirth [22]. However, women are less likely to be identified or treated as they demonstrate fewer signs and symptoms of NCD, for instance, CVD, compared to men and get a lack of attention in case of disease prevention strategies [25].

Although Bangladesh has been successful in achieving several Sustainable Development Goals (SDG), the government and many international organizations are still working to decrease the burden of NCD as these are considered a challenge in the field of public health. It is evident that to prevent and control NCD, the socio-demographic determinants of major NCD risk factors and their clustering should be explored and understood in a systematic way. There are many studies that focused on the prevalence and risk factors of NCDs [1,5–7,10,11], in addition to diabetes, hypertension [1,13,14], overweight and obesity [12]. Though a large number of studies were nationally representative studies and few studies used earlier data or focused on a specific geographic region, studies on the women of reproductive age addressing the overall social determinants and prevalence of risk factors of NCD were very rare. This study, therefore, aimed to investigate the prevalence of risk factors of NCD and determine their socio-demographic distributions among Bangladeshi reproductive-aged women.

## Materials and Methods

### Study population and data source

This study used the data from the latest Bangladesh Demographic and Health Survey (BDHS) 2017-2018 to explore the prevalence of non-communicable disease risk factors among reproductive-aged women. The BDHS is a nationally representative cross-sectional survey and covers the entire population residing in non-institutional dwelling units in the country. The BDHS survey was performed using a stratified, two-stage household sample. In the first step, 675 enumeration areas (EAs) (250 in urban areas and 425 in rural areas) were picked with a probability proportional to the size of the EA. In the second round of sampling, an average of 30 households per EA were gathered independently from urban and rural areas and from each of the eight divisions in order to obtain statistically reliable estimates of key demographic and health factors for the entire country. Detailed survey sampling and the data collection procedure have been published in the BDHS survey report [26]. On the survey, 20,250 households were selected, interviews were completed in 19,457 households with an overall 96.5% household response rate, and information was recorded from 89,819 household members [26]. Finally, this study only included 5,624 reproductive-aged women (age 18-49 years) who had their blood pressure, height and weight, and smoking status recorded.

### Outcome variables

The outcome variables for this research were the three risk factors for non-communicable diseases, which are smoking, overweight and obesity, and hypertension. These three selected NCD risk factors are available in the BDHS 2017-2018 survey data set.

#### Smoking

In the BDHS survey, participants’ current smoking status was recorded as a dichotomous response.

#### Overweight and obesity

Height and weight measures were collected for respondents in all of the households sampled. Weight measurements were obtained using an electronic scale (model number SECA 878U) with a digital screen, and height measurements were carried out with a measuring board (ShorrBoard). The health technician and one female technician were deployed to take both measurements. Women who were pregnant on the day of the survey visit or had given birth during the preceding two months were excluded. Body mass index (BMI) was calculated by dividing weight in kilograms by height in meters squared (kg/m^2^). The overweight was considered for the individual with BMI between 25.0 and 29.9, and obese was BMI greater than or equal to 30.0 [26].

#### Hypertension

Blood pressure was measured for all participants aged 18 and above in the subsample of one-fourth of the households. All participants who qualified for blood pressure measures were contacted and instructed on the procedure. Those who gave their consent were measured for their blood pressure. The automatic device includes separate cuffs BP monitor was used to measuring the blood pressure. Three blood pressure measurements were taken at intervals of approximately 10 minutes. Typically, the average of the second and third measurements was utilized to reflect the blood pressure results of respondents. Individuals were diagnosed having hypertension if they had an average systolic blood pressure level of 140 mmHg or above, and diastolic blood pressure level of 90 mmHg or above, or they were taking antihypertensive medication at the time of the survey [27,28].

### Explanatory variables

This study considered the socio-demographic covariates related to smoking, overweight/obesity, and hypertension. These included the respondents’ ages (18–29, 30–39, and 40–49 years), education (no education, primary, secondary, and higher-level), marital status (married, never married, and widowed/divorced), residence (rural, urban), geographical areas (Barishal, Chittagong, Dhaka, Khulna, Mymensingh, Rajshahi, Rangpur, and Sylhet divisions), occupational status (not working, service, agriculture/self-employed, and manual), wealth index (poorest, poorer, middle, richer and richest). The wealth index of the households was calculated by the principal component analysis. The scores were given based on available consumer goods in the household (television, bicycle, car, etc.), and household characteristics (source of drinking water, sanitation facilities, flooring materials, etc.). Then the national wealth quintiles were compiled by assigning the household score to each usual household member, ranking each person in the household population by her/his score, and then dividing the distribution into five equal categories, each comprising 20% of the population [26].

### Data analysis

The socio-demographic characteristics of the respondents were reported in mean and weighted percentages. The crude prevalence of smoking, overweight and obesity, and hypertension was determined after accounting for the complex survey design and survey sampling weights. Differences between categorical variables were tested using the chi-square tests. We applied mixed-effects Poisson regression models with robust variance to identify factors linked with non-communicable diseases risk factors and reported the results as adjusted prevalence ratio (APR) with a 95% confidence interval (CI). We used this model since the odds ratio estimated using logistic regression from a cross-sectional study may significantly overestimate relative risk when the outcome is common [29,30]. Secondly, in the case of convergence failure with the log-binomial model, Poisson regression with a robust variance performs better in estimating the prevalence ratio from a cross-sectional study [31]. Finally, the number of risk factors present within each participant (from 0 to 3) was counted to assess the clustering of risk factors and analyzed using the mixed-effects Poisson regression. All statistical tests were two-tailed, and a p-value of <0.05 was regarded as statistically significant. Statistical software STATA-16 (Stata Corp LP, College Station, TX, USA) was used to conduct the analysis.

### Ethical considerations

This study used secondary data from the country representative survey (BDHS 2017-2018). The study protocol for BDHS was approved by the ICF (international institutional review board), and the data is publicly available (http://dhsprogram.com/data/available-datasets.cfm). Therefore, no further ethical approval was necessary for this study. However, we received authorization from the DHS to use the datasets. Informed consent was obtained from each participant of the survey before enrolling in the survey by using the Introduction and Consent form of the survey. It was also explained that the information will be kept strictly confidential and will not be shared with anyone except with the members of the survey team.

## Results

### Socio-demographic characteristics of study participants

The socio-demographic characteristics of the study participants (n=5,624) are shown in **Table 1**. The mean age of research participants was 31 years (SD=9.07). Approximately half (48.26%) of them were between 18 and 29 years old. In terms of educational attainment, 17.78% of the participants did not have any formal education. On the other hand, only 14.67% of respondents have education above the secondary level. In this study, 88.22% of respondents were married. More than two-thirds (71.84%) of the participants resided in rural areas. However, about one-quarter (24.44%) of the participants came from the Dhaka division, which is the capital city of Bangladesh. The richest comprised 22.02% of the total participants based on the wealth index. Surprisingly, the proportions of participants in between the poorest and richer quintiles were almost identical. More than half (54.38%) of the participants were not involved in any formal work.

**Table 1.** Socio-demographic characteristics of participants.

| **Variables** | **Un-weighted count** | **Weighted percent/Mean (SD)** |
| --- | --- | --- |
| **Total** | 5,624 | 100.00 |
| **Mean age (year)** |  | 31.00 (9.07) |
| **Age group (year)** |  |  |
| 18 – 29 | 2,689 | 48.26 |
| 30 – 39 | 1,716 | 30.26 |
| 40 – 49 | 1,219 | 21.48 |
| **Educational status** |  |  |
| No education | 957 | 17.78 |
| Primary | 1,691 | 29.87 |
| Secondary | 2,051 | 37.68 |
| Higher | 925 | 14.67 |
| **Marital status** |  |  |
| Never married | 402 | 6.33 |
| Married | 4,902 | 88.22 |
| Widowed/divorced | 320 | 5.45 |
| **Residence** |  |  |
| Rural | 3,533 | 71.84 |
| Urban | 2,091 | 28.16 |
| **Division** |  |  |
| Barishal | 590 | 5.47 |
| Chattogram | 827 | 18.57 |
| Dhaka | 799 | 24.44 |
| Khulna | 729 | 11.69 |
| Mymensingh | 589 | 7.50 |
| Rajshahi | 732 | 14.24 |
| Rangpur | 690 | 11.62 |
| Sylhet | 668 | 6.46 |
| **Wealth index** |  |  |
| Poorest | 1,069 | 19.01 |
| Poorer | 1,051 | 19.50 |
| Middle | 1,081 | 19.90 |
| Richer | 1,091 | 19.56 |
| Richest | 1,332 | 22.02 |
| **Occupational status\*** |  |  |
| Not working | 3,066 | 54.38 |
| Services | 242 | 4.01 |
| Agriculture/self-employed | 1,667 | 30.22 |
| Manual | 641 | 11.39 |

\*8 cases missing

### Prevalence of non-communicable diseases risk factors among reproductive-aged women

**Table 2** depicts the prevalence of risk factors for non-communicable diseases among reproductive-aged women. The prevalence of smoking was 9.55% among the study participants. Smoking was more prevalent among the age group 40-49 years (19.26%), women with no educational status (19.69%), Sylhet division residents (21.69%), participants with the poorest wealth index (14.04%), and the agriculture/self-employed women (14.49%) (p <0.001). Compared to other risk factors, overweight and obesity were profound among the participants (31.57%). About 40.26% of the participants between 30-39 years were either overweight or obese (p <0.001). Likewise, overweight and obesity were more prevalent among married (33.36%), urban residents (40.18%), Chattogram division residents (36.26%), the richest (52.31%), and service holders (35.18%) (p <0.001). In this study, 20.27% of women were suffering from hypertension. Women aged 40 to 49 had the highest prevalence of hypertension (39.21%) (p <0.001). In addition, the prevalence of hypertension was higher among women who were uneducated (29.07%), and divorced/widowed (25.70%) (p <0.001).

**Table 2.** Prevalence of non-communicable diseases risk factors among reproductive-aged women.

| Variables | Current smoking |  | Overweight and obesity |  | Hypertension |  |
| --- | --- | --- | --- | --- | --- | --- |
|  | n | Prevalence, % (95% CI) | n | Prevalence, % (95% CI) | n | Prevalence, % (95% CI) |
| **Total** | 5,624 | 9.55 (8.65-10.53) | 5603 | 31.57 (30.17-33.02) | 5624 | 20.27 (19.11-21.49) |
| **Age group (year)** |  |  |  |  |  |  |
| 18 – 29 | 2,689 | 4.28 (3.53-5.17) | 2,679 | 22.92 (21.11-24.82) | 2,689 | 9.03 (7.87-10.33) |
| 30 – 39 | 1,716 | 11.06 (9.48 -12.86) | 1,708 | 40.26 (37.63-42.94) | 1,716 | 24.77 (22.48-27.20) |
| 40 – 49 | 1,219 | 19.26 (16.69 -22.12) | 1,216 | 38.82 (35.91-41.82) | 1,219 | 39.21 (36.16-42.35) |
| *P-value* |  | <0.001 |  | <0.001 |  | <0.001 |
| **Educational status** |  |  |  |  |  |  |
| No education | 957 | 19.69 (17.05-22.62) | 952 | 26.13 (23.26-29.21) | 957 | 29.07 (25.92-32.43) |
| Primary | 1,691 | 12.58 (10.95-14.41) | 1,688 | 30.75 (28.35-33.27) | 1,691 | 22.29 (20.16-24.57) |
| Secondary | 2,051 | 5.07 (4.17-6.15) | 2,044 | 33.81 (31.42-36.29) | 2,051 | 17.14 (15.39-19.04) |
| Higher | 925 | 2.60 (1.67-4.05) | 919 | 34.08 (30.44-37.92) | 925 | 13.57 (11.33-16.17) |
| *P-value* |  | <0.001 |  | 0.001 |  | <0.001 |
| **Marital status** |  |  |  |  |  |  |
| Never married | 402 | 1.39 (0.53-3.57) | 396 | 10.43 (7.47-14.38) | 402 | 6.18 (4.01-9.40) |
| Married | 4,902 | 9.71 (8.78-10.73) | 4,890 | 33.36 (31.88-34.87) | 4,902 | 20.95 (19.69-22.27) |
| Widowed/divorced | 320 | 16.41 (2.42-21.38) | 317 | 26.85 (21.57-32.87) | 320 | 25.70 (20.60-31.56) |
| *P-value* |  | <0.001 |  | <0.001 |  | <0.001 |
| **Residence** |  |  |  |  |  |  |
| Rural | 3,533 | 10.21 (9.11-11.42) | 3,519 | 28.20 (26.53-29.94) | 3,533 | 19.92 (18.50-21.41) |
| Urban | 2,091 | 7.87 (6.45-9.57) | 2,084 | 40.18 (37.63-42.78) | 2,091 | 21.19 (19.21-23.31) |
| *P-value* |  | 0.024 |  | <0.001 |  | 0.317 |
| **Division** |  |  |  |  |  |  |
| Barishal | 590 | 21.29 (17.65-25.45) | 588 | 29.28 (25.13-33.80) | 590 | 24.24 (20.43-28.50) |
| Chattogram | 827 | 8.62 (6.64-11.10) | 824 | 36.26 (32.57-40.11) | 827 | 23.51 (20.71-26.57) |
| Dhaka | 799 | 6.35 (4.77-8.41) | 795 | 35.73 (32.44-39.16) | 799 | 16.90 (14.37-19.78) |
| Khulna | 729 | 6.83 (4.77-9.70) | 728 | 35.45 (31.80-39.29) | 729 | 20.47 (17.29-24.05) |
| Mymensingh | 589 | 17.01 (13.72-20.89) | 587 | 24.69 (21.03-28.75) | 589 | 15.76 (12.94-19.06) |
| Rajshahi | 732 | 5.06 (3.24-7.82) | 728 | 28.13 (24.64-31.90) | 732 | 20.62 (17.36-24.32) |
| Rangpur | 690 | 8.92 (6.10-12.86) | 689 | 25.20 (21.89-28.83) | 690 | 23.36 (20.51-26.48) |
| Sylhet | 668 | 21.69 (17.73-26.24) | 664 | 24.40 (20.11-29.26) | 668 | 18.93 (15.78-22.55) |
| *P-value* |  | <0.001 |  | <0.001 |  | 0.001 |
| **Wealth index** |  |  |  |  |  |  |
| Poorest | 1,069 | 14.04 (11.72-16.75) | 1,066 | 16.48 (14.10-19.19) | 1,069 | 17.35 (14.77-20.28) |
| Poorer | 1,051 | 12.46 (10.43-14.81) | 1,046 | 21.74 (19.12-24.62) | 1,051 | 18.84 (16.44-21.48) |
| Middle | 1,081 | 8.37 (6.76-10.31) | 1,080 | 29.80 (26.82-32.97) | 1,081 | 20.10 (17.66-22.78) |
| Richer | 1,091 | 8.42 (6.80-10.39) | 1,088 | 34.59 (31.47-37.84) | 1,091 | 21.39 (18.82-24.21) |
| Richest | 1,332 | 5.16 (3.98-6.66) | 1,323 | 52.31 (49.24-55.37) | 1,332 | 23.24 (20.81-25.86) |
| *P-value* |  | <0.001 |  | <0.001 |  | 0.019 |
| **Occupational status\*** |  |  |  |  |  |  |
| Not working | 3,066 | 7.53 (6.53-8.67) | 3,052 | 34.35 (32.41-36.34) | 3,066 | 20.19 (18.65-21.83) |
| Services | 242 | 5.62 (3.15-9.85) | 238 | 35.18 (28.54-42.45) | 242 | 20.76 (15.51-27.20) |
| Agriculture/self-employed | 1,667 | 14.49 (12.57-16.63) | 1,667 | 25.97 (23.85-28.20) | 1,667 | 22.25 (20.03-24.63) |
| Manual | 641 | 7.60 (5.33-10.72) | 638 | 32.46 (28.52-36.68) | 641 | 15.56 (12.83-18.75) |
| *P-value* |  | <0.001 |  | <0.001 |  | 0.013 |

\*8 cases missing

Furthermore, the research determined the prevalence of NCD risk factors by the number of factors across different age groups (**Fig. 2**). More than one-third of the participants had one NCD risk factor, and 12.51% of participants had two NCD risk factors.

**Fig 1.**
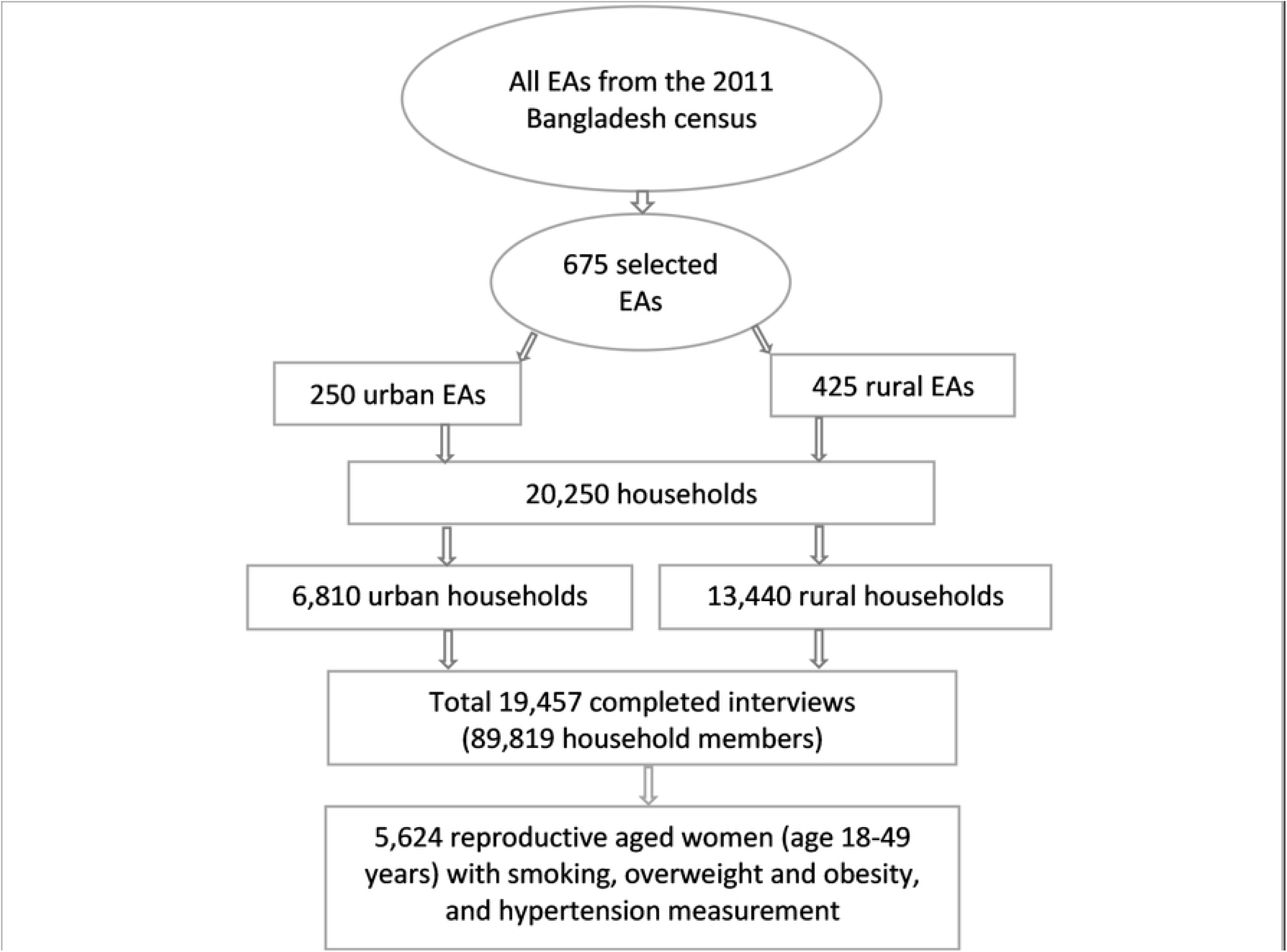
Schematic representation of the sampling procedure of the Bangladesh Demographic and Health Survey, 2017–2018.

**Fig. 2.**
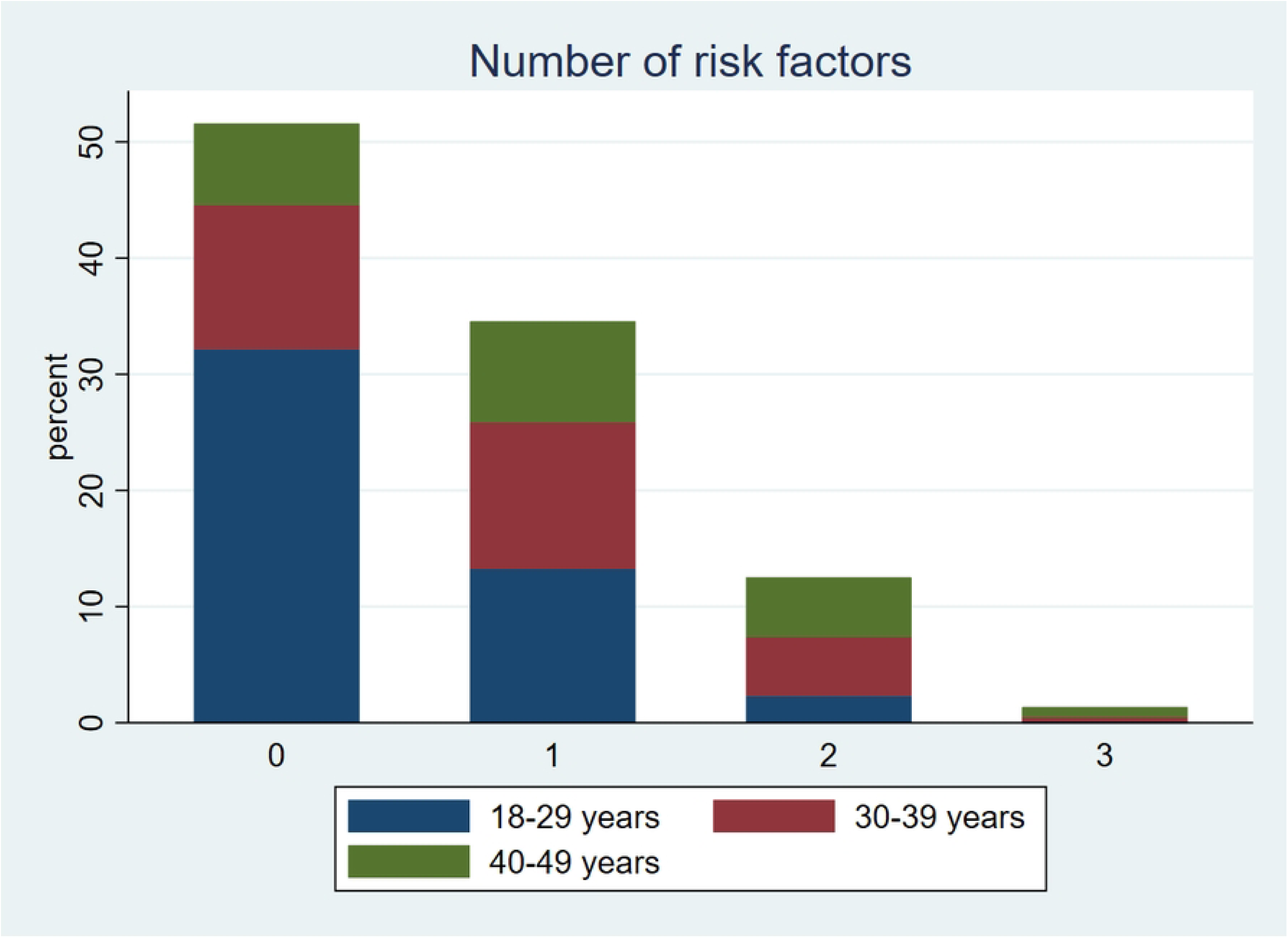
Prevalence of number of non-communicable diseases risk factors among reproductive-aged women.

### Association between socio-demographic characteristics and non-communicable diseases risks factors

According to **Table 3**, the mixed-effects Poisson regression model revealed the adjusted association between socio-demographic characteristics and non-communicable diseases risk factors. The participants’ age was strongly associated with the risk factors for non-communicable diseases. Compared to participants aged 18-29 years, those aged 30-39 years (APR: 1.83; 95% CI: 1.45-2.30) and 40-49 years (APR: 2.65; 95% CI: 2.07-3.38) were more likely to be smokers. The prevalence ratio for overweight and obesity in the age group 30-39 years (APR: 1.71; 95% CI: 1.55-1.89) and 40-49 years (APR: 1.77; 95% CI: 1.57-1.98) were significantly higher than the age group18-29 years. Similarly, the prevalence of hypertension was significantly higher in the 30-39 years (APR: 2.58; 95% CI: 2.18-3.05) and 40-49 years (APR: 3.94; 95% CI: 3.34-4.69) age groups compared to the younger age group. The prevalence ratio of smoking was almost 3 times higher among participants with no education (APR: 3.19; 95% CI: 1.85-5.49) compared to those with a higher level of education. Married women (APR: 2.71; 95% CI: 1.96-3.75) and widowed/divorced women (APR: 2.16; 95% CI: 1.49-3.15) demonstrated a higher prevalence of overweight and obesity compared to the never-married women.

**Table 3.**
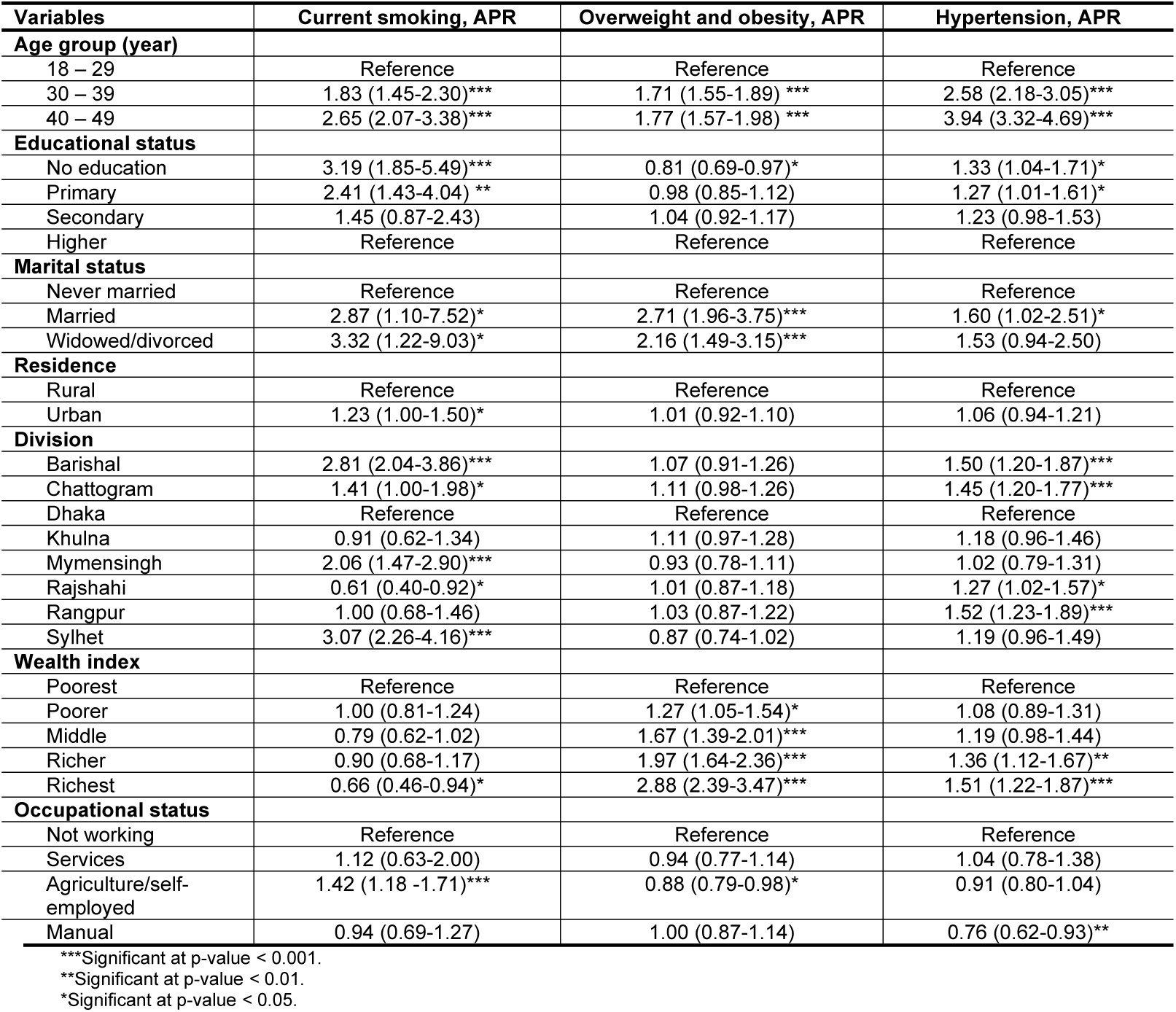
Association between socio-demographic characteristics and non-communicable diseases risk factors identified from the mixed-effects Poisson regression model.

| Variables | Current smoking, APR | Overweight and obesity, APR | Hypertension, APR |
| --- | --- | --- | --- |
| **Age group (year)** |  |  |  |
| 18 – 29 | Reference | Reference | Reference |
| 30 – 39 | 1.83 (1.45-2.30)*** | 1.71 (1.55-1.89) *** | 2.58 (2.18-3.05)*** |
| 40 – 49 | 2.65 (2.07-3.38)*** | 1.77 (1.57-1.98) *** | 3.94 (3.32-4.69)*** |
| **Educational status** |  |  |  |
| No education | 3.19 (1.85-5.49)*** | 0.81 (0.69-0.97)* | 1.33 (1.04-1.71)* |
| Primary | 2.41 (1.43-4.04) ** | 0.98 (0.85-1.12) | 1.27 (1.01-1.61)* |
| Secondary | 1.45 (0.87-2.43) | 1.04 (0.92-1.17) | 1.23 (0.98-1.53) |
| Higher | Reference | Reference | Reference |
| **Marital status** |  |  |  |
| Never married | Reference | Reference | Reference |
| Married | 2.87 (1.10-7.52)* | 2.71 (1.96-3.75)*** | 1.60 (1.02-2.51)* |
| Widowed/divorced | 3.32 (1.22-9.03)* | 2.16 (1.49-3.15)*** | 1.53 (0.94-2.50) |
| **Residence** |  |  |  |
| Rural | Reference | Reference | Reference |
| Urban | 1.23 (1.00-1.50)* | 1.01 (0.92-1.10) | 1.06 (0.94-1.21) |
| **Division** |  |  |  |
| Barishal | 2.81 (2.04-3.86)*** | 1.07 (0.91-1.26) | 1.50 (1.20-1.87)*** |
| Chattogram | 1.41 (1.00-1.98)* | 1.11 (0.98-1.26) | 1.45 (1.20-1.77)*** |
| Dhaka | Reference | Reference | Reference |
| Khulna | 0.91 (0.62-1.34) | 1.11 (0.97-1.28) | 1.18 (0.96-1.46) |
| Mymensingh | 2.06 (1.47-2.90)*** | 0.93 (0.78-1.11) | 1.02 (0.79-1.31) |
| Rajshahi | 0.61 (0.40-0.92)* | 1.01 (0.87-1.18) | 1.27 (1.02-1.57)* |
| Rangpur | 1.00 (0.68-1.46) | 1.03 (0.87-1.22) | 1.52 (1.23-1.89)*** |
| Sylhet | 3.07 (2.26-4.16)*** | 0.87 (0.74-1.02) | 1.19 (0.96-1.49) |
| **Wealth index** |  |  |  |
| Poorest | Reference | Reference | Reference |
| Poorer | 1.00 (0.81-1.24) | 1.27 (1.05-1.54)* | 1.08 (0.89-1.31) |
| Middle | 0.79 (0.62-1.02) | 1.67 (1.39-2.01)*** | 1.19 (0.98-1.44) |
| Richer | 0.90 (0.68-1.17) | 1.97 (1.64-2.36)*** | 1.36 (1.12-1.67)** |
| Richest | 0.66 (0.46-0.94)* | 2.88 (2.39-3.47)*** | 1.51 (1.22-1.87)*** |
| **Occupational status** |  |  |  |
| Not working | Reference | Reference | Reference |
| Services | 1.12 (0.63-2.00) | 0.94 (0.77-1.14) | 1.04 (0.78-1.38) |
| Agriculture/self-employed | 1.42 (1.18 -1.71)*** | 0.88 (0.79-0.98)* | 0.91 (0.80-1.04) |
| Manual | 0.94 (0.69-1.27) | 1.00 (0.87-1.14) | 0.76 (0.62-0.93)** |

\*\*\*Significant at p-value < 0.001.
\*\*Significant at p-value < 0.01.
\*Significant at p-value < 0.05.

For current smoking, prevalence ratio of the participants from Barishal (APR: 2.81; 95% CI: 2.04-3.86), Mymensingh (APR: 2.06; 95% CI: 1.47-2.90), and Sylhet (APR: 3.07; 95% CI: 2.26-4.16) was comparatively higher than the participants from Dhaka, the capital of Bangladesh. However, hypertension was almost 1.5 times higher among the participants from Barishal, Chattogram, and Rangpur than the participants from Dhaka. In addition, individuals with hypertension (APR: 1.51; 95% CI: 1.22-1.87), overweight and obesity (APR: 2.88; 95% CI: 2.39-3.47) were more likely to belong to the wealthiest quintile. When compared to the group of people who were not employed, those whose occupations involved agriculture or self-employment had a higher prevalence ratio for current smoking (APR: 1.42; 95% CI: 1.18-1.71).

### Results of mean number NCD risk factors and multivariable analysis of clustering of NCD risk factors

We have examined the clustering of NCD risk factors (the number of risk factors present in an individual from 0 to 3) in our sample. **Table 4** shows that women of 40–49 years were more likely to experience non-communicable diseases risk factors than 18–29 years aged women (ARR: 2.44; 95% CI: 2.22-2.68). Women with no education were at slightly higher risk for NCD risk factors (ARR: 1.15; 95% CI: 1.00-1.33) compared to the women with a higher level of education. In comparison with the never-married women, married (ARR: 2.32; 95% CI: 1.78–3.04), and widowed/divorced (ARR: 2.14; 95% CI: 1.59–2.89) were more prevalent in NCD risk factors.

**Table 4.** Mean number of NCD risk factors and multivariable analysis (mixed-effects Poisson regression model) of clustering of NCD risk factors.

| Variables | Mean number (95% CI) | Clustering of NCD risk factors, APR |
| --- | --- | --- |
| **Age group (year)** |  |  |
| 18 – 29 | 0.36 (0.34-0.39) | Reference |
| 30 – 39 | 0.76 (0.72-0.80) | 1.94 (1.78-2.11)*** |
| 40 – 49 | 0.97 (0.92-1.03) | 2.44 (2.22-2.68)*** |
| **Educational status** |  |  |
| No education | 0.75 (0.69-0.80) | 1.15 (1.00-1.33)* |
| Primary | 0.66 (0.61-0.70) | 1.13 (1.00-1.29)* |
| Secondary | 0.56 (0.53-0.60) | 1.09 (0.97-1.23) |
| Higher | 0.50 (0.45-0.55) | Reference |
| **Marital status** |  |  |
| Never married | 0.18 (0.13-0.23) | Reference |
| Married | 0.64 (0.62-0.66) | 2.32 (1.78-3.04)*** |
| Widowed/divorced | 0.69 (0.59-0.78) | 2.14 (1.59-2.89)*** |
| **Residence** |  |  |
| Rural | 0.58 (0.56-0.61) | Reference |
| Urban | 0.69 (0.65-0.73) | 1.06 (0.99-1.14) |
| **Division** |  |  |
| Barishal | 0.75 (0.68-0.82) | 1.44 (1.28-1.63)*** |
| Chattogram | 0.68 (0.63-0.74) | 1.25 (1.12-1.40)*** |
| Dhaka | 0.59 (0.53-0.64) | Reference |
| Khulna | 0.63 (0.57-0.69) | 1.10 (0.97-1.24) |
| Mymensingh | 0.58 (0.51-0.64) | 1.13 (0.98-1.30) |
| Rajshahi | 0.54 (0.48-0.60) | 1.02 (0.90-1.16) |
| Rangpur | 0.58 (0.52-0.63) | 1.15 (1.01-1.32)* |
| Sylhet | 0.65 (0.58-0.72) | 1.27 (1.12-1.43)*** |
| **Wealth index** |  |  |
| Poorest | 0.48 (0.43-0.53) | Reference |
| Poorer | 0.53 (0.48-0.58) | 1.11 (0.99-1.26) |
| Middle | 0.58 (0.54-0.63) | 1.25 (1.11-1.41)*** |
| Richer | 0.64 (0.59-0.69) | 1.44 (1.27-1.63)*** |
| Richest | 0.81 (0.76-0.85) | 1.82 (1.60-2.07)*** |
| **Occupational status** |  |  |
| Not working | 0.62 (0.59-0.65) | Reference |
| Services | 0.62 (0.50-0.73) | 0.99 (0.83-1.18) |
| Agriculture/self-employed | 0.63 (0.59-0.67) | 0.98 (0.90-1.06) |
| Manual | 0.55 (0.49-0.61) | 0.90 (0.80-1.01) |

\*\*\*Significant at p-value < 0.001.
\*\*Significant at p-value < 0.01.
\*Significant at p-value < 0.05.

Individuals in the Barishal (ARR: 1.44; 95% CI: 1.28–1.63), Chattogram (ARR: 1.25; 95% CI: 1.12–1.40), and Sylhet (ARR: 1.27; 95% CI: 1.12–1.43) divisions were living with somewhat higher risk factors of NCD compared to the individuals in Dhaka division. Likewise, women belonged to the middle (ARR: 1.25; 95% CI: 1.11–1.41), richer (ARR: 1.44; 95% CI: 1.27–1.63), and richest (ARR: 1.82; 95% CI: 1.60–2.07) were more likely to have the risk factors of NCD compared to the poorest wealth quintile. Furthermore, the regression prevalence ratio plot also demonstrates the determinants of clustering of NCD risk factors (**Fig. 3**).

**Fig. 3.**
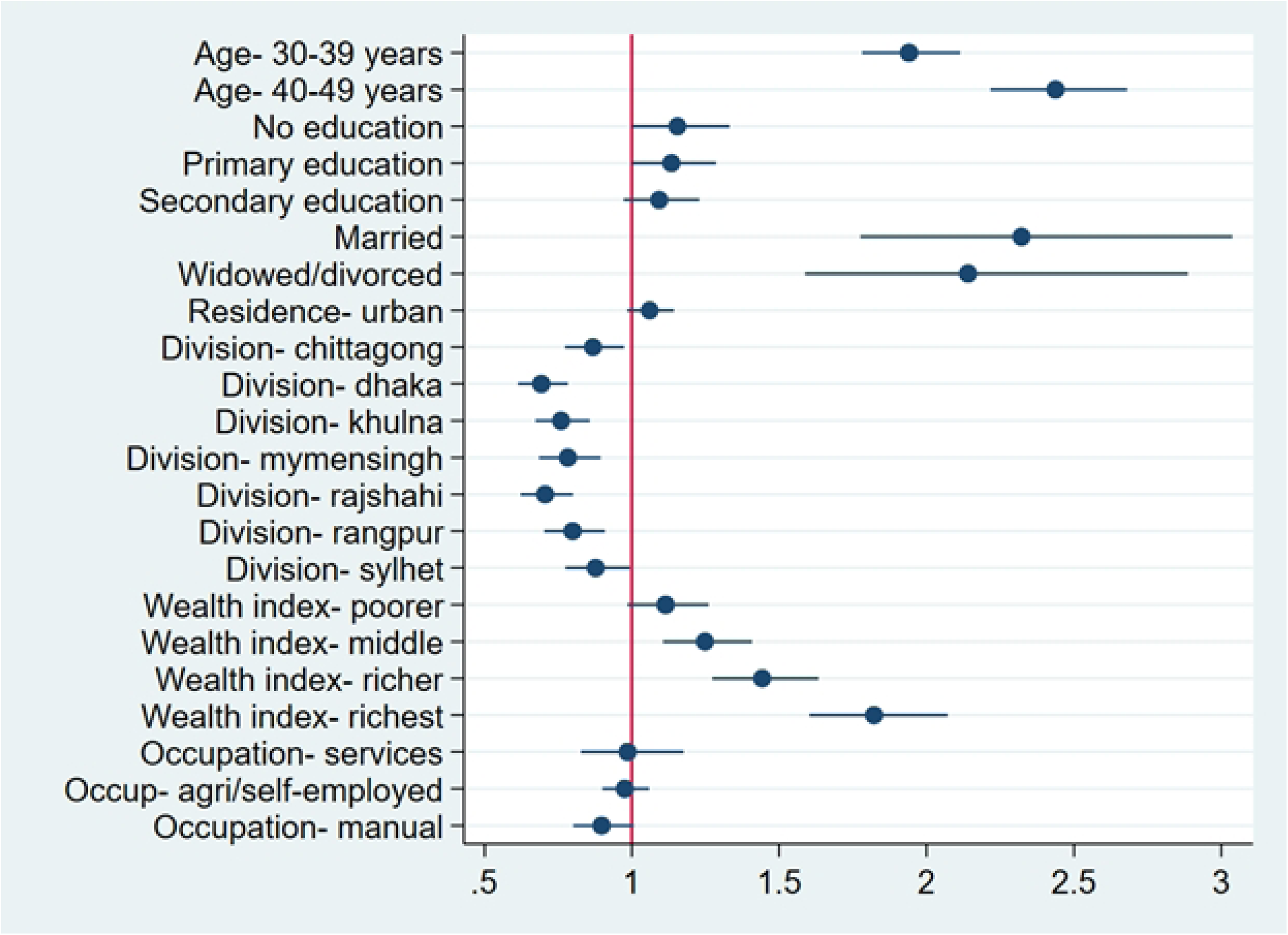
Regression prevalence ratio plot for the socio-demographic characteristics associated with NCD risk factors identified from the mixed-effects Poisson regression model.

## Discussion

It is well established that NCD are increasing at an alarming rate [2,3]. Globally, the prevalence of NCD was reported much higher in women in comparison to men [2]. The scenario is not different for Bangladesh as well [32]. It has been reported that in recent years, the prevalence of NCD doubled in reproductive-aged women of LMICs [33]. So, our study attempted to investigate the prevalence and determinants of NCD risk factors among reproductive-aged women of Bangladesh by using the most recent nationally representative survey.

We found that the smoking, overweight/obesity, and hypertension rates were 9.55%, 31.57%, and 20.27%, respectively, among reproductive-aged women. Our findings were similar to other previous study findings of Bangladesh that also reported similar rates of NCD risk factors [1,32,34,35]. The result of our study showed that the prevalence of smoking increased with age. The findings of our study were in line with the findings of a similar nationwide survey held in Nepal [36]. The high prevalence of smokers was reported commonly among women aged 40+ years, living in a rural area, illiterate, widowed/divorced, and with lower socio-economic status, which is similar to other studies [36–39]. Similar to previous studies, a three-fold higher prevalence of smoking among aged (30-40 years), divorced, and illiterate might be a reflection of coping strategies, and smoking might be an option to overcome their loneliness [40]. Bista et al. found in their study in Nepal that the proportion of tobacco usage was more than 2 times higher among 30-40 years aged women and nearly four times higher among 40+years women in comparison to 15-29 years women [36]. The trend was similar for our study as well. This might be due to the cultural and socio-demographic similarities between Nepal and Bangladesh. Sreeramareddy et al. found similar results while comparing nationwide surveys in nine South-Asian countries, including Bangladesh and Nepal [41]. Respondents from urban areas reported being more smokers. This result is similar to the previous study conducted in Bangladesh [42]. Also, we have found that participants from Sylhet had reported more than 3 times higher smoking rates than Dhaka (capital). Similar area level variation was reported in a previous study conducted in Bangladesh [43].

We observed an increasing trend in overweight/obesity and hypertension with increasing age and wealth index, similar to Bista et al. [36]. Though hypertension steadily increased with age, the prevalence of overweight and obesity inconsistently increased with age. Women in the 30-39 years age group were reported to be more obese (40.26%), whereas 40-49 years aged women had the highest rate of hypertension (39.21%). However, the prevalence of overweight and obesity was twice in both aforementioned age groups compared to the 18-29 years age group, which is aligned with the findings of Khanam et al. [44]. The increasing trend of hypertension with age is also evident in another nationwide study in Bangladesh [45]. We observed education level has vice versa relation with hypertension and overweight & obesity. The hypertension rate was higher in the illiterate group, whereas the obesity rate was higher in people who had secondary education. Similar findings were described by Sun K. et al., who explained that participants with lower educational backgrounds have to involve in more physically active work than educated persons, which explained their lower rate of overweight and obesity [46]. This result also can be explained by the fact that the participants from the agriculture/self-employed group had a lower prevalence of overweight and obesity in our study. Obesity and hypertension both can endanger women’s life during pregnancy, so these alarming conditions need to be addressed and should intervene policies.

We found that married women were more vulnerable to hypertension and prone to be obese than never-married women. Findings were supported by previous studies [36,44]. Taking birth control methods might be an explanation for these findings [47,48]. Similar to other nationwide studies, we found that wealth index and geographical areas had a significant role in overweight/obesity and hypertension [44,45]. Obesity and hypertension both significantly increased with the wealth index. This positive association with the wealth index is a clear reflection of less physical activity due to their sedentary lifestyle. Though there was no significant difference between the residential distribution for both obesity and hypertension, similar to Chowdhury et al., we found that the highest hypertensive participants were reported from Rangpur, and the lowest was from Sylhet. Poverty, malnutrition, and salt intake patterns might be responsible for this variation [49,50].

We have also found that nearly 13% of the study population reported having two risk factors, which was similar to the findings of Biswas et al. [35]. We have seen that the clustering of NCD risk factors increased with age and wealth index. The finding of our study was aligned with the trend of increasing NCD risk factors found in previous studies from different countries, including Bangladesh [35,36]. Our study demonstrates that lower education is a potential confounder for NCD risk factors. However, it was inconsistent with the findings from other population-based studies of Bangladesh [11,51]. A specific study population with a specific age range might be responded differently from existing studies that resulted in the distribution of NCD risk factors among the respondents. Urbanization had no significant impact on NCD risk factors, though, in participants from Barishal, NCD risk factors were reported as approximately 1.5 times more than in Dhaka. Findings are similar to the research of Al-Zubayer et. al. [51].

Bangladesh has achieved the Millennium Development Goal-5 (MDG-5) [52]; however, the findings of this study demonstrate that reproductive-aged women are still in a vulnerable position. These findings indicate an alarming situation for NCD among reproductive-aged women in Bangladesh. It might be a barrier for us to achieving Sustainable Development Goal (SDG) [53]. Preexisting NCD risk factors are well-established determinants for several adverse health outcomes, including maternal death [54]. Many adverse pregnancy outcomes are significantly associated with obesity, hypertension, and the smoking status of mothers [55–57]. Considering the age of our study participants, these factors might be responsible for doubling the risk of unfavorable health conditions.

### Strength and Limitation

This study is the first study in Bangladesh that investigated prevalence and determinants of NCD risk factors among reproductive-aged women. The major strength of our study is that it used a population-based, nationally representative data source. The BDHS covered rural and urban regions in all administrative divisions, making these findings generalizable to the country. Our mixed-effects Poisson regression corrects the overestimation of the effects size produced by conventional logistic regression employed in cross-sectional studies and increases the precision of the findings. Like the other studies, we had some limitations as well. As we used a secondary data source, we were only allowed to find out the associations between the dependent and independent variables. We failed to show any causal relationship. Secondly, the BDHS did not record other important clinical biomarkers (i.e., blood lipid profile, HbA1c, S. Creatinine), as well as behavioral factors (e.g., alcohol consumption, sleep duration), dietary factors (e.g., type and amount of food taken), physical activity, which are crucial risk factors for NCD. So, these data could not be included in the analyses, which limits the strength of this study.

## Conclusions

There was a high prevalence of NCD risk factors among older women and who were currently married or widowed/divorced, from the wealthiest socio-economic groups, and with less education. Furthermore, the relationship between NCD risk factors and geographic location was found to be significant. Lessening the burden of non-communicable diseases necessitates effective disease prevention and control measures for women through reducing the burden of hypertension, overweight/obesity, and smoking. If the government does not address these risk factors, women of reproductive age in Bangladesh will be at risk for poor health outcomes and it will be difficult to reach the sustainable development goal of reducing non-communicable diseases.

## Data Availability

All relevant data are within the manuscript and its Supporting Information files.

## Acknowledgments

The authors thank MEASURE DHS (Demography and Health Surveys) for granting access to the BDHS 2017–2018 data.

## Supporting information

Dataset of the study can be found in **S1 Data** file.

